# Effect of intravenous lidocaine dose on pain upon awakening in pediatric tonsillectomy: A randomized trial

**DOI:** 10.1101/2024.06.21.24309278

**Authors:** Yang Hu, Ming-Cheng Du, Yi Chen, Xiang Long, Jing-Jing Jiang, Yuan Gong

## Abstract

We investigated the potential of intravenous lidocaine to reduce pain on awakening in a dose-dependent manner and aimed to determine the median effective dose of lidocaine in 200 children aged 3–12 years (American Society of Anesthesiologists physical status I–II) who underwent elective tonsillectomy with or without adenoidectomy at Yichang Central People’s Hospital. The patients were randomized into four dose groups (A–D: 0.0, 1.0, 1.5, and 2.0 mg/kg, respectively), and they underwent the same anesthetic induction and maintenance protocols. The primary outcome was pain on awakening, while secondary outcomes included pain scores at 1, 4, 12, and 24 h after awakening; incidences of laryngospasm, bronchospasm, or perioperative stridor; and time to extubation. Intraoperative cardiac events were classified as safety events. Our findings indicated that intravenous lidocaine dose-dependently reduced pain on awakening, with the median effective dose being 1.75 mg/kg. Significant differences were observed between groups A and D (P ≤ 0.001). No incidents of laryngospasm, bronchospasm, or perioperative stridor were observed. Furthermore, there were significant between-group differences in time to extubation (P ≤ 0.05). In conclusion, our study demonstrated that lidocaine dose-dependently reduced pain on awakening in children undergoing tonsillectomy with or without adenoidectomy, with no severe adverse events.

## Introduction

Tonsillectomy, with or without adenoidectomy, is one of the most common surgical procedures performed in children wherein children receive general anesthesia via a tracheal tube [1,2]. Although this surgery is a common and safe procedure in children, severe pain on awakening is frequently observed following pediatric adenotonsillectomy [3]. It can easily turn into persistent pain without appropriate treatment and drastically limits patients’ oral intake of fluids and pain medication, potentially extending postoperative length of stay [4,5].

Lidocaine is a local anesthetic that is used as a general anesthetic adjunct because of its analgesic, anti-hyperalgesic, anti-inflammatory, and immunomodulatory properties related to stress [6–8]. Intravenous lidocaine reduces pain after elective surgery under general anesthesia [9]. However, it is unclear whether lidocaine reduces pain on awakening in a dose-dependent manner, and the median effective dose (ED_50_) of intravenous lidocaine in children remains unknown.

We hypothesized that intravenous lidocaine reduces pain on awakening in a dose-dependent manner and determined the ED_50_ of lidocaine in children undergoing tonsillectomy with or without adenoidectomy. The choice of pain on awakening as the primary outcome was guided by the need for clinically meaningful outcomes with potential benefits.

## Materials and methods

### Ethical Considerations

All procedures performed in studies involving human participants were in accordance with the ethical standards of the institutional and/or national research committee and with the 1964 Declaration of Helsinki and its later amendments or comparable ethical standards. The study design was approved by the Institutional Review Board (president, Ke-Jun Yan) of Yichang Central People’s Hospital (approval number: HEC-KYJJ-2020-038-02; approval date: September 17, 2021). Written informed consent was obtained from the parents or guardians of all children enrolled in the study.

### Study Design

This single-center, parallel-group, double-blind, randomized controlled trial (principal investigator, Yuan Gong) was registered in the Chinese Clinical Trial Registry (http://chictr.org.cn/showproj.aspx?proj=136605) (registration number: ChiCTR2100053006; registration date: November 8, 2021). The trial was conducted from December 1, 2021, to May 30, 2022, at Yichang Central People’s Hospital, Hubei Province, China, in accordance with the CONSORT guidelines. Patients were randomized to each group in an equal (1:1:1:1) ratio. Randomization was computer-generated, and each patient was assigned a code.

### Patients

The inclusion criteria were patients aged 3–12 years (American Society of Anesthesiologists physical status I–II) who were scheduled for elective tonsillectomy with or without adenoidectomy. Children were divided into the following four groups, according to the dose of lidocaine administered: A (0.0 mg/kg), B (1.0 mg/kg), C (1.5 mg/kg), and D (2.0 mg/kg). The exclusion criteria were chronic cough, a history of steroid or bronchodilator treatment, reactive airway disease, upper airway infection 2 weeks before the procedure, angiotensin-converting enzyme inhibitor therapy, gastroesophageal reflux, morbid obesity, known allergy to any of the study drugs, use of medications or nutraceuticals known to affect blood pressure (BP) and heart rate (HR), surgery lasting > 2 h, unexpected bleeding, and the need for > 2 intubation attempts.

### Perioperative Anesthetic Care

Preoperatively, all children fasted for 6 h and were restricted from oral intake of clear fluids for 1 h. The children entered the operating room accompanied by their parents to curb separation anxiety. Non-invasive BP, HR, and pulse oxygen saturation were measured, and electrocardiography was performed using a multifunction monitor (GE Healthcare, Chicago, IL, USA). The width of the BP cuff for each patient was approximately two-thirds of the length of the upper arm. After stabilization for 5 min, baseline HR, systolic BP, diastolic BP, and mean arterial pressure values were obtained from the average of three measurements taken 2 min apart. A 22-gauge intravenous catheter was subsequently inserted into a vein on the back of the hand.

After preoxygenation, the drug of interest (lidocaine [Anhui Changjiang Pharmaceutical Co. Ltd., Wuhu City, China] or 0.9% saline) was administered intravenously for 3 s. An anesthetic nurse blinded to the study prepared the drug of interest to be administered through an infusion pump. General anesthesia was induced 2 min after administration, using the following induction protocol: sufentanil (0.25 μg/kg [Yichang Renfu Pharmaceutical Co. Ltd., Yichang City, China]), propofol (2.0 mg/kg [Fresenius Kabi Deutschland GmbH, Homburg, Germany]), and rocuronium (0.6 mg/kg [Zhejiang Xianju Pharmaceutical Co. Ltd., Taizhou City, Zhejiang Province, China]). An anesthetist blinded to the study graded the cough response during injection. When the eyelash reflex disappeared, the lungs were ventilated via a face mask with 100% oxygen. A cuffed tracheal tube was used, the size of which was selected based on a widely used formula (3.5 + [age (years)] / 4). The patient was excluded from the study if any difficulty was encountered while performing face mask ventilation. Anesthesia was maintained with 2–3% sevoflurane (Maruishi Pharmaceutical Co. Ltd., Osaka, Japan) and 50% oxygen.

At the end of the procedure, sevoflurane was discontinued, and neostigmine (0.04 mg/kg [Zhejiang Xianju Pharmaceutical Co. Ltd.]) and atropine (0.02 mg/kg [Suicheng Pharmaceutical Co. Ltd., Xinzheng City, China]) were administered to antagonize any residual neuromuscular blockade. Following surgery, oral suction was immediately performed, with the patient still under anesthesia. Extubation was performed while the patient recovered after confirming an adequate tidal volume, a regular spontaneous respiratory pattern, and purposeful behavior (eyes opened on request). After extubation, an anesthetist who was not involved in the study assessed recovery from anesthesia, scored the throat pain, and graded the cough response. Patients were monitored for ≥ 5 min, with 100% oxygen via a face mask, to allow regular spontaneous respiration. Patients were transferred to a post-anesthesia care unit (PACU) after extubation. The time to extubation (from sevoflurane discontinuation to tracheal extubation) was recorded. Electrocardiography, peripheral pulse oximetry, and non-invasive BP measurements were also performed.

Patients were discharged from the PACU when their Steward score was > 4. Other postoperative care was performed according to the practices of local clinicians. If the pain (Face, Legs, Activity, Cry, and Consolability scale [children aged 3–4 years], Wong-Baker scale [children aged 4–7 years], or visual analog scale [children aged ≥8 years])^3^ score was ≥ 3 at rest, the attending PACU nurse administered intravenous propacetamol (30 mg/kg) as treatment. The use of other medications was restricted.

### Primary Outcome

The primary outcome was pain on awakening.

### Secondary Outcomes

The secondary outcomes were pain scores at 1, 4, 12, and 24 h after awakening; incidences of laryngospasm, bronchospasm, and perioperative stridor; and time to extubation. Safety events included intraoperative cardiac events (arrhythmia, hypotension [mean arterial pressure < 65 mmHg], hypertension [mean arterial pressure > 90 mmHg]), and rescue treatment.

### Statistical analyses

The sample size was calculated based on an expected pain on-awakening score difference of 1 (clinical significance), with 80% power (α = 0.05, β = 0.2), which indicated that 50 patients were required per group. Patient characteristics (including age, height, and weight); pain on awakening scores; incidences of laryngospasm, bronchospasm, and perioperative stridor; duration of surgery; and time to extubation were expressed as mean ± SD and analyzed using ANOVA, with a P-value < 0.05 indicating a significant difference. Statistical significance was set at P < 0.01. All statistical analyses were conducted using GraphPad Prism (version 8.0.2; GraphPad Software Inc., San Diego, CA, USA).

## Results

### Patients

Between December 1, 2021, and May 30, 2022, 200 patients were enrolled and divided into four groups based on the dose of lidocaine administered(Fig. 1). There were significant differences in the time to extubation between groups A and B (P = 0.014), A and C (P = 0.015), and A and D (P = 0.017). Patient characteristics and other operative data were not significantly different between the groups (Table 1).

**Fig. 1.**
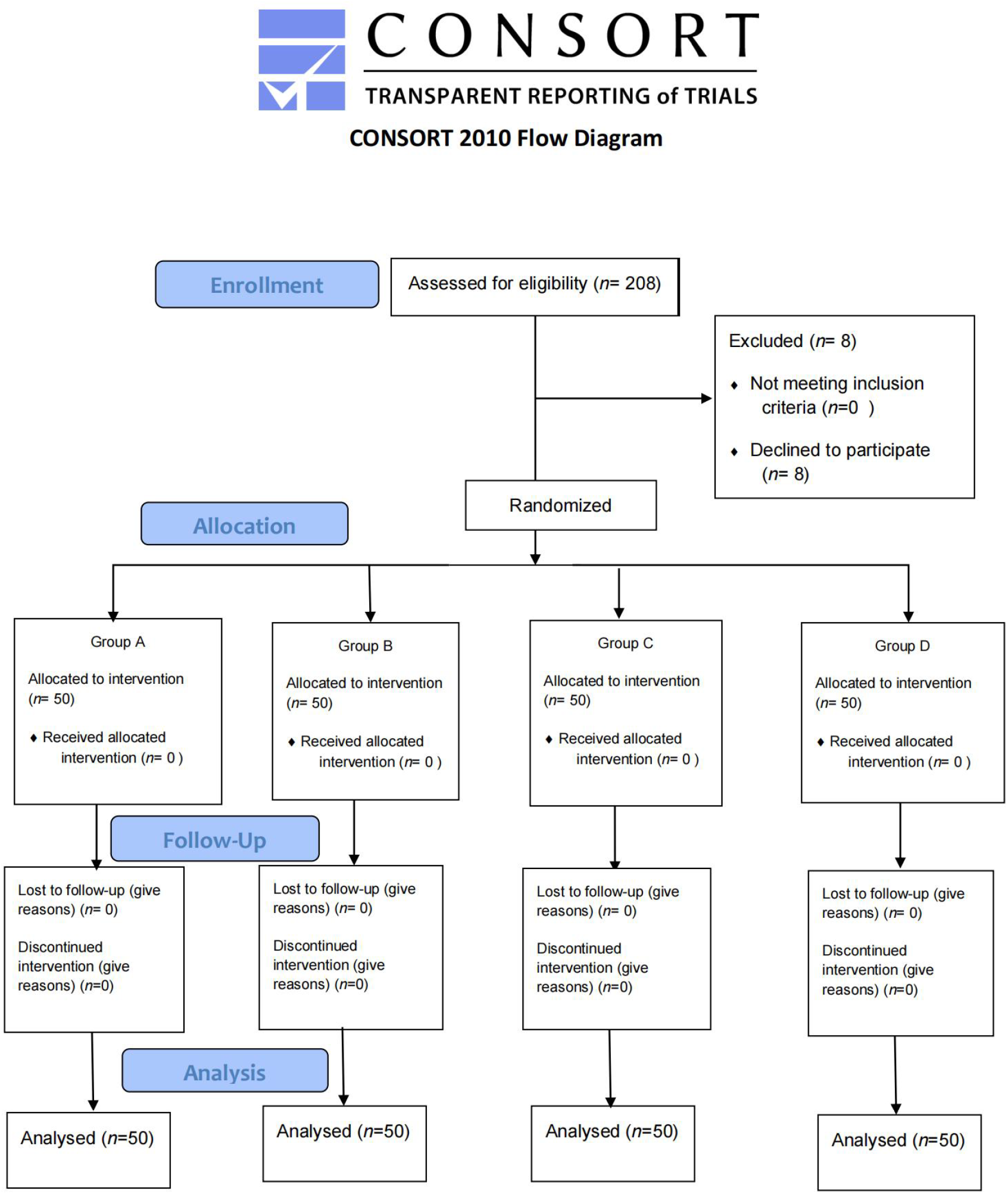
Flow Diagram

**Table 1.**
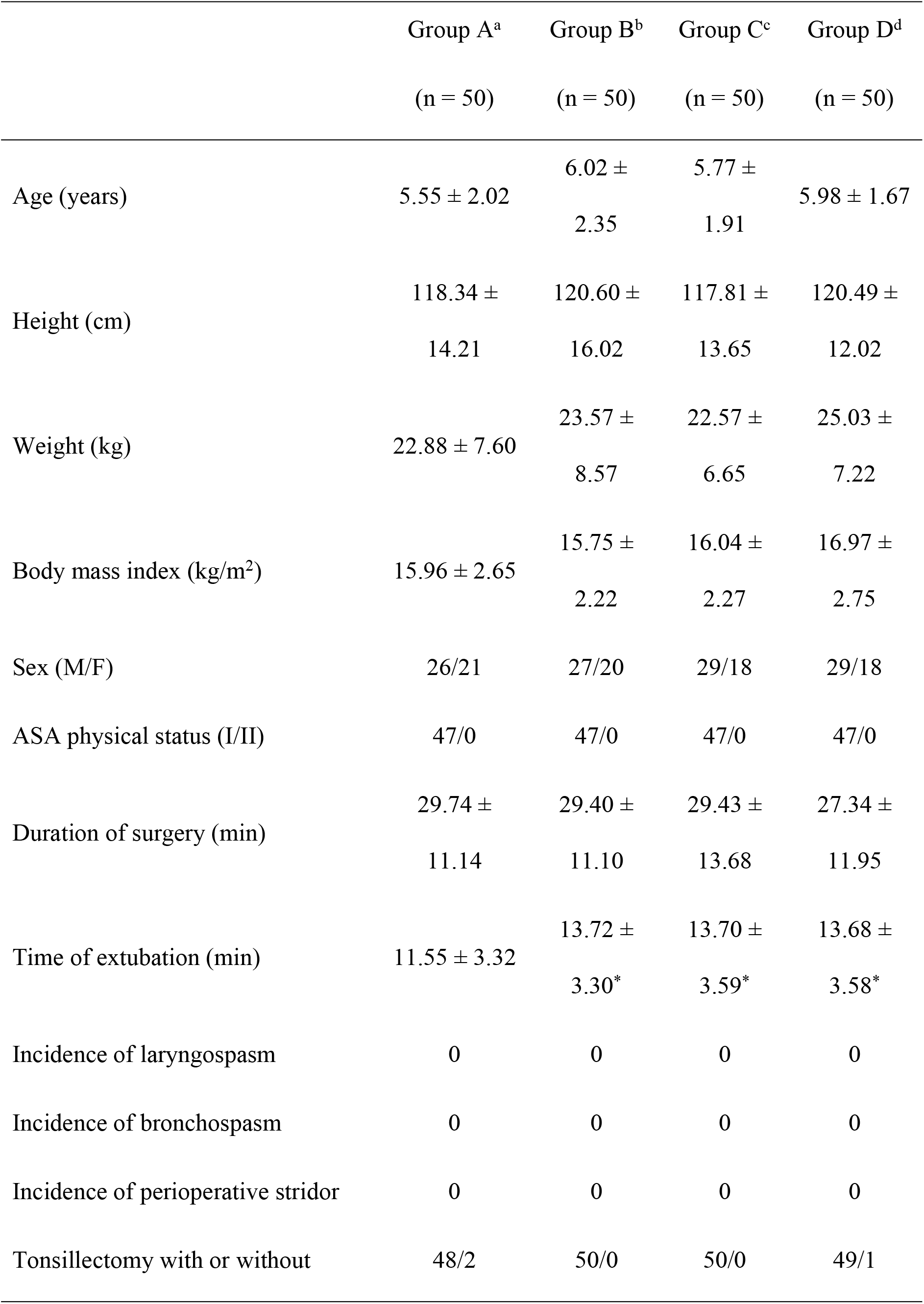
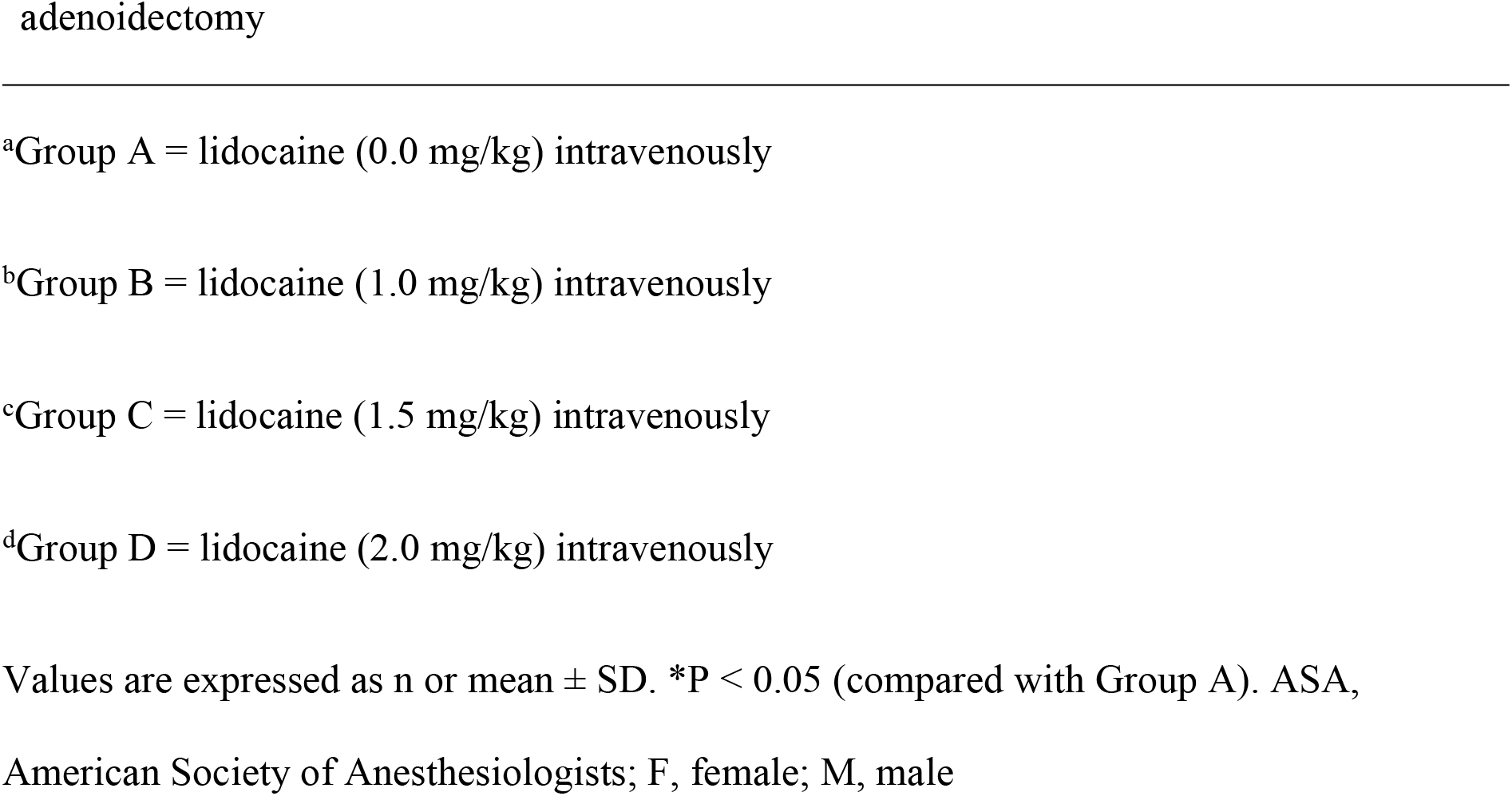
Patient characteristics and clinical data.

### Primary outcomes

The ED_50_ of intravenous lidocaine for pain on awakening was 1.75 mg/kg (Fig. 2a). All 50 patients in each group were assessed for the primary outcome. The score for pain on awakening was significantly different between the groups, in a dose-dependent manner, with significant differences between groups C and D (P = 0.037), A and B (P = 0.007), B and D (P = 0.003), and A and C (P ≤ 0.001). There was also a significant difference between groups A and D (P ≤ 0.001) (Fig. 2b).

**Fig. 2.**
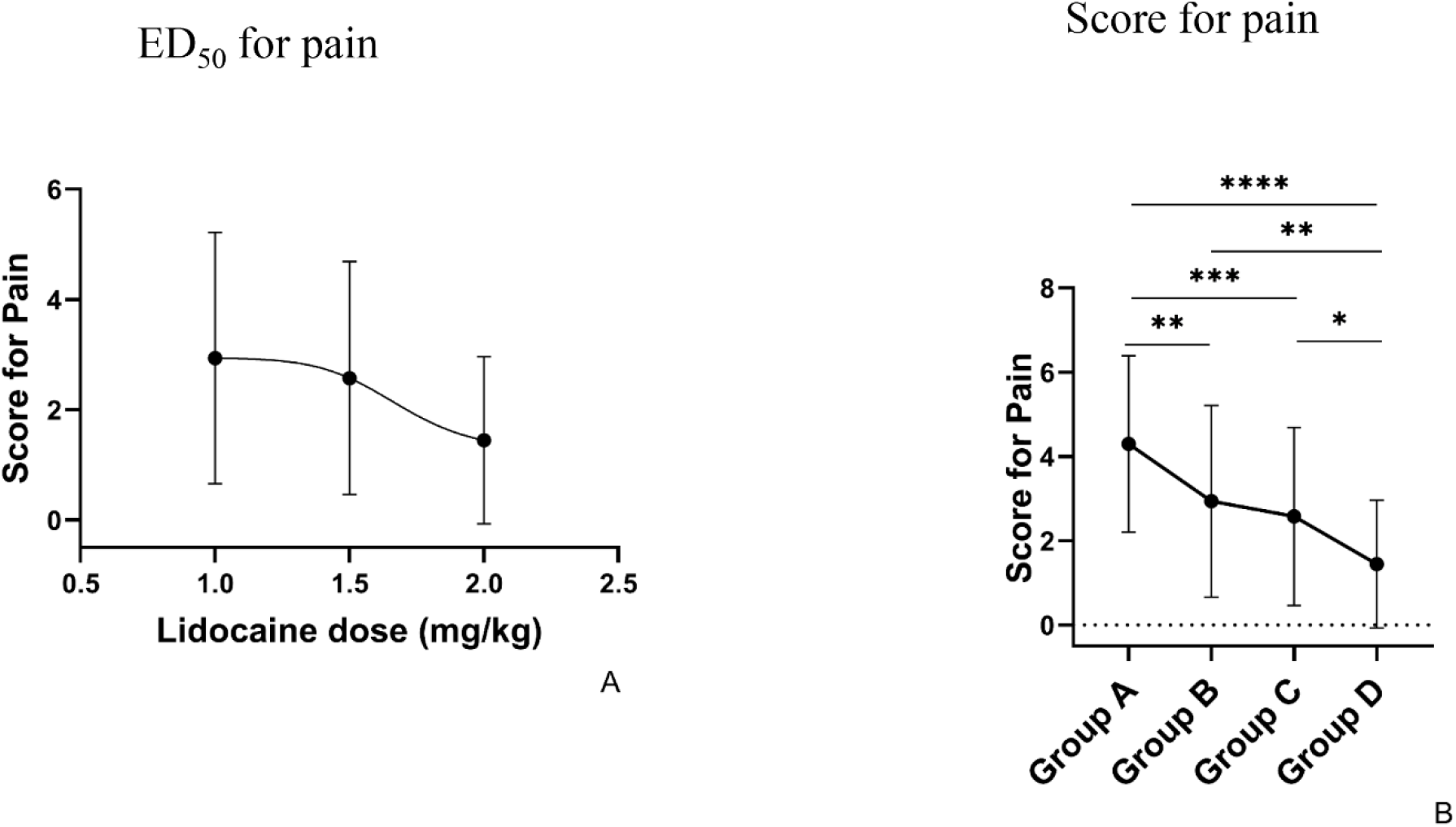
Intravenous lidocaine for pain on awakening. (a) Concentration-response curve. (b) Pain on awakening scores. ^*^*P* < 0.05; ^**^*P* < 0.01; ^***^*P* < 0.001; ^****^*P* < 0.0001. ED_50_, median effective dose.

### Secondary outcomes

The pain score at 1 h after awakening was significantly different between groups A and B (P ≤ 0.001); A, C, and D (P ≤ 0.001); and B and D (P = 0.010). The pain score at 4 h after awakening was significantly different between groups A and B (P ≤ 0.001); A, C, and D (P ≤ 0.001); and B and D (P = 0.015); however, there were no significant differences in the pain scores at 12 and 24 h after awakening (Fig. 3). No cases of laryngospasm, bronchospasm, or perioperative stridor were observed in any group. However, significant differences in the time to extubation were observed between groups A and B (P = 0.014), A and C (P = 0.015), and A and D (P = 0.017) (Table 1). No severe complications, such as arrhythmia, were observed in any group.

**Fig. 3.**
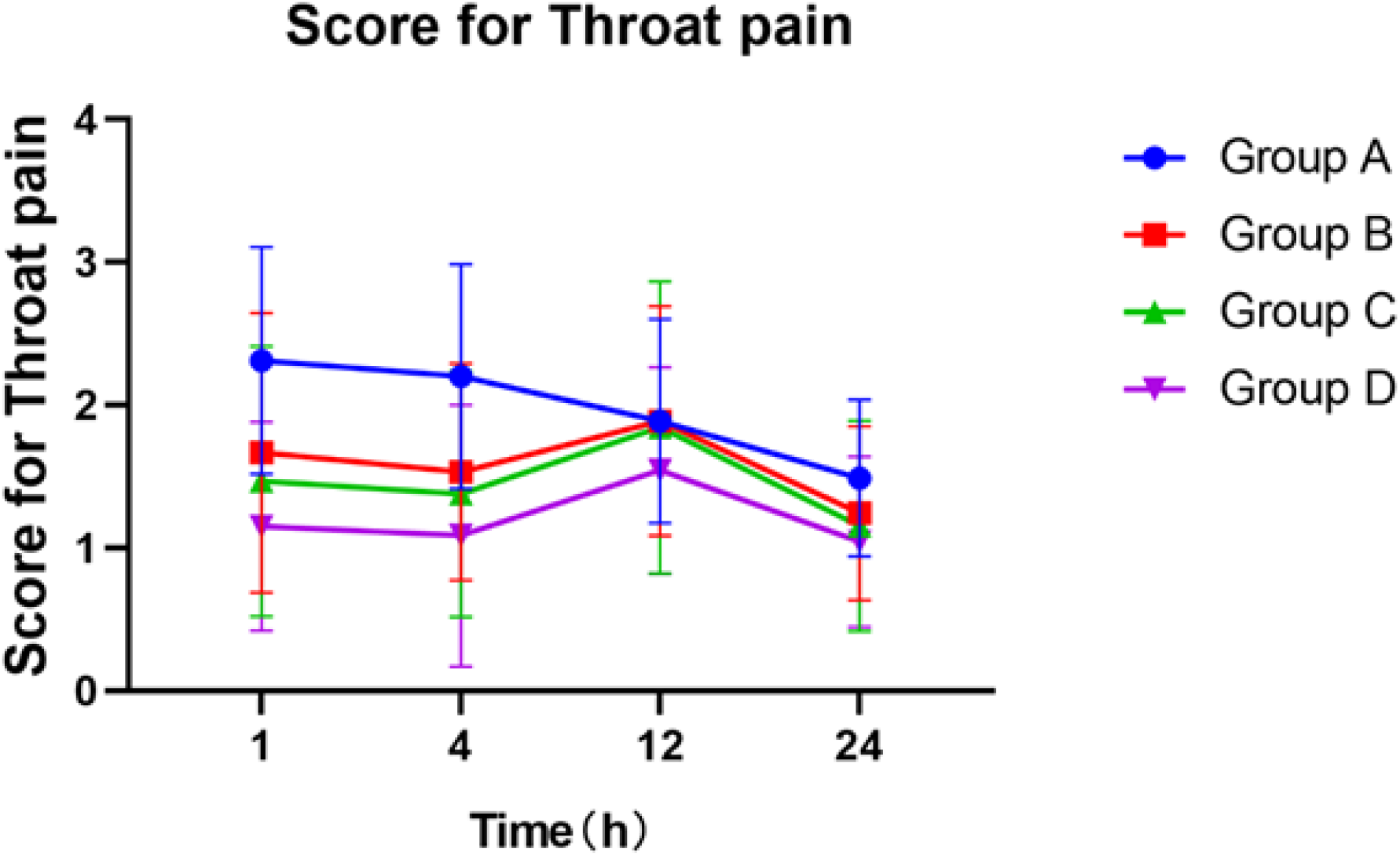
Pain scores at 1, 4, 12, and 24 h after awakening.

## Discussion

Tonsillectomy is one of the most painful surgical procedures. The relief of pain after tonsillectomy remains challenging and controversial. Several interventions have been recommended to control pain after tonsillectomy in children [10]. Opioids are an important treatment strategy for postoperative pain. However, there is a tendency to avoid using narcotics for respiratory depression in children. Large studies of non-steroidal anti-inflammatory drugs have demonstrated effective pain management [10]. However, their potential for treatment of more severe bleeding events has been debated.

In this trial, intravenous lidocaine resulted in a dose-dependent reduction in pain on awakening, with an ED_50_ of 1.75 mg/kg. Even 1 and 4 h after surgery, there was a considerable reduction in pain scores for those who received lidocaine compared with those who did not receive lidocaine. Only after 12 h did the pain scores on awakening exhibit no difference between groups. This seems feasible as the duration of surgery was < 1 h and the half-life of intravenous lidocaine was approximately 2 h [11]. The exact inhibitory mechanism of lidocaine is unclear. It may involve its analgesic, anti-hyperalgesic, anti-inflammatory, and immunomodulatory properties related to stress [6–8].

No cases of laryngospasm, bronchospasm, perioperative stridor, or other serious complications were observed during anesthesia. Intravenous lidocaine appears safe and does not result in serious adverse events in children. Substantial differences in the time to extubation were observed between the study (B, C, and D) groups and the control (A) group; the time in the control group was 2 min longer to meet extubation requirements.

This study has several limitations. First, it was a single-center study, which reduced its power and reliability. Second, we used the sevoflurane concentration in end-expiratory gas to monitor the depth of anesthesia, potentially leading to an increase in the ED_50_.

## Conclusion

Intravenous lidocaine reduced pain on awakening in children undergoing tonsillectomy with or without adenoidectomy in a dose-dependent manner without causing serious adverse events. The ED_50_ of intravenous lidocaine for pain on awakening was 1.75 mg/kg.

## Data Availability

Data are available from the Ethics Committee (contact via corresponding author) for researchers who meet the criteria for access to confidential data.

## Acknowledgments

We would like to thank Editage (www.editage.com) for English language editing.

